# Thoracic spine mobilization on autonomic nervous system in a healthy population - a randomized controlled double-blinded feasibility study

**DOI:** 10.64898/2026.02.28.26347312

**Authors:** Slavko Rogan, Gerard Farrell, Susanne Schlarb, Marcus Schlarb, Shagun Agarwal, Ron Clijsen

**Affiliations:** Bern University of Applied Sciences, School of Health Professions, Discipline Physiotherapy, Bern, Switzerland; University of Otago, Centre for Health, Activity, and Rehabilitation Research, School of Physiotherapy, Dunedin, New Zealand; Osteopathie Schlarb, Geretsried, Germany; Galgotias University, School of Allied Health Sciences, Department of Physiotherapy, Greater Noida, India; Vrije Universiteit Brussel, Department of Movement and Sport Sciences, Faculty of Physical Education and Physiotherapy, Brussels, Belgium; University of Applied Sciences and Arts of Southern Switzerland, Department of Business Economics, Health and Social Care, Rehabilitation and Exercise Science Laboratory (RESlab), Landquart, Switzerland

**Keywords:** thoracic spine mobilization, autonomic nervous system, heart rate variability, feasibility study

## Abstract

**Background:** Thoracic spine mobilization (TSM) has been proposed to influence autonomic nervous system (ANS) activity, yet evidence remains inconsistent and feasibility of standardised protocols is unclear. This study aimed to evaluate whether a randomized TSM protocol can be implemented successfully in healthy participants and to provide preliminary estimates of its effects on heart rate variability (HRV) and heart rate (HR).

**Methods:** A randomized feasibility trial was conducted with healthy young adults receiving six manual therapy sessions consisting of rotational mobilizations above Th5 over 14 days. Feasibility outcomes included adherence, absence of unexpected adverse events (UAE), and practicality of autonomic data acquisition. Physiological outcomes comprised HRV parameters, high-frequency (HF), low-frequency/high-frequency ratio (LF/HF) and HR, analyzed using autoregressive (AR) and fast Fourier transform (FFT) methods.

**Results:** Procedural safety and methodological integrity were confirmed (no UAE; complete datasets), but feasibility was only partially achieved due to adherence shortfalls, higher attrition, and device-related delays. Physiologically, large effect sizes were observed in the intervention group: at evening assessment, HF_AR showed ES = 0.80 (p = .008); at morning assessment, HF_FFT ES = 0.72 (p = .016), HF_AR ES = 0.78 (p = .010), and LF/HF_AR ES = 0.70 (p = .021). HR remained unchanged. These findings suggest repeated TSM may modulate HRV, primarily through HF-related changes associated with vagal activity, while LF/HF interpretation remains controversial.

**Conclusion:** A randomized TSM protocol is safe and methodologically viable with logistical refinements. Preliminary evidence indicates potential vagal modulation, warranting larger trials with respiratory control, ECG-based HRV, multimodal ANS measures, and clinical populations to confirm efficacy and translational relevance.

## Background

The autonomic nervous system (ANS), comprising two branches, the sympathetic nervous system (SNS) and parasympathetic nervous system (PNS), regulates cardiovascular function, tissue trophicity, and pain modulation. Traditionally, physiotherapists assume thoracic spine mobilization (TSM) reduces SNS activity, while cervical techniques influence vagal tone. However, this anatomical view is overly simplistic. As Neuhuber (2009) and Jänig (2022) emphasize, the SNS innervates the entire body, whereas the PNS supplies only specific regions; thus, the popular notion of a global antagonism between SNS and PNS is misleading. Organ control via dual innervation is the exception rather than the rule, and functional analysis reveals a complex interplay rather than a simple opposition. This system-level perspective suggests that research should focus on how manual therapy influences ANS activity overall, rather than assuming predictable segmental effects.

Evidence on TSM and sympathetic–parasympathetic balance (SPB) remains limited. A scoping review by Rogan et al. (2023) found that 74% of studies reported ANS responses after spinal manual therapy, with 51% showing increased SNS activity and 24% increased PNS activity, independent of treatment region. Similarly, Hansen et al. (2025) concluded that thoracic mobilization can affect both SNS and PNS markers in symptomatic and asymptomatic participants. These findings align with neurophysiological models (Bialosky et al., 2009; Rogan et al., 2019; Slavko Rogan et al., 2016) that integrate peripheral and central pathways, including midbrain circuits such as the periaqueductal gray (PAG), which modulates cardiovascular and nociceptive responses (Lovick, 1985).

Manual therapy may act through two pathways: a biomechanical mechanism improving joint mobility and a physiological mechanism involving mechano-cellular signalling and reflex-mediated changes (Rogan et al., 2019; Rogan et al., 2017). Animal studies show that PAG stimulation increases HR, BP, and respiratory rate alongside SNS activation and hypoalgesia, supporting central involvement in autonomic modulation (Lovick, 1985). Human studies report mixed results: Slater et al.(1994) observed SNS changes after sympathetic slump mobilization; Zegarra-Parodi et al. (2016) found increased skin blood flow (interpreted as SNS inhibition); Vicenzino et al. (1998) and McGuiness et al. (1997) reported HR and BP increases after cervical mobilization; while Sampath et al. (2017) found no SNS changes after thoracic manipulation. HRV changes have also been documented (Giles et al., 2013; Younes et al., 2017; Zhang et al., 2006), but inconsistencies persist.

Most studies measure HRV at artificially constructed time points, such as scheduled visits or follow-ups, which risks producing stylized data that do not reflect real-life autonomic regulation. To improve ecological validity, HRV measurement should be simplified and integrated into participants’ normal routines. This study adopts this innovative approach by assessing HRV responses to TSM in a practical setting rather than under highly controlled conditions. The main goal was to evaluate feasibility and explore whether repeated TSM sessions can influence ANS activity in healthy individuals.

ANS activity was assessed using HRV, BP, HR, and segmental skin perfusion and erythema. HRV frequency analysis distinguishes HF (≈0.15–0.40 Hz; vagal) and LF (≈0.04–0.15 Hz; mixed sympathetic-parasympathetic); LF/HF ratio is used cautiously as an SPB index (Chouchou et al., 2013; Lansdown & Rees, 2012). HRV and BP are validated autonomic markers (Dreifus et al., 1993; Younes et al., 2017). Skin perfusion and erythema provide proxies for cutaneous vasomotor activity.

Randomized controlled trials (RCTs) offer strong causal inference but are resource-intensive (Evans, 2003; Oakley et al., 2006). Feasibility studies are essential to determine whether recruitment, intervention delivery, and multimodal autonomic measurements can be implemented successfully before launching a full-scale trial (Eldridge, Lancaster, et al., 2016; Rogan & Karstens, 2018; Thabane et al., 2010). To date, only one feasibility study has investigated the effects of TSM on level T1–T5 (Tal et al., 2018) and at levels T6-T12 (Rogan et al., 2019) on HRV, BP, and HR. Building on the systems-level ANS framework (Jänig, 2022; Neuhuber, 2009) and region-independent effects reported by Rogan (2023) and Hansen et al. (2025), this study aimed to evaluate feasibility and provide preliminary estimates of TSM effects on SPB. Therefore, the present feasibility study aimed primarily to evaluate whether a randomized protocol for TSM can be implemented successfully in healthy participants. Feasibility was assessed by participant adherence, the absence of UAE, and the practicality of data acquisition and analysis for autonomic markers. The secondary aim was to provide preliminary estimates of the effects of six sessions consisting of rotational mobilizations of the thoracic spine above Th5 on ANS outcomes, measured via HRV and HR. Specifically, the research question was: What changes occur in HRV and related autonomic parameters following six manual therapy sessions consisting of rotational mobilizations of the thoracic spine above Th5 within 14 days? These findings will inform the design and sample size calculation for future randomized controlled trials.

## Methods

### Study design

This feasibility study included healthy adults aged 20 to 35 years who were randomly allocated to an intervention group or a control group (Figure 1). The age range was selected to minimize confounding from autonomic maturation, which continues until approximately age 20, and from age related endocrine decline beginning around age 35. Both processes influence autonomic regulation (Farrell et al., 2023). A control group rather than a placebo group was chosen because the primary aim was to assess feasibility outcomes such as recruitment, adherence, acceptability, safety, and data acquisition.

**Figure 1:**
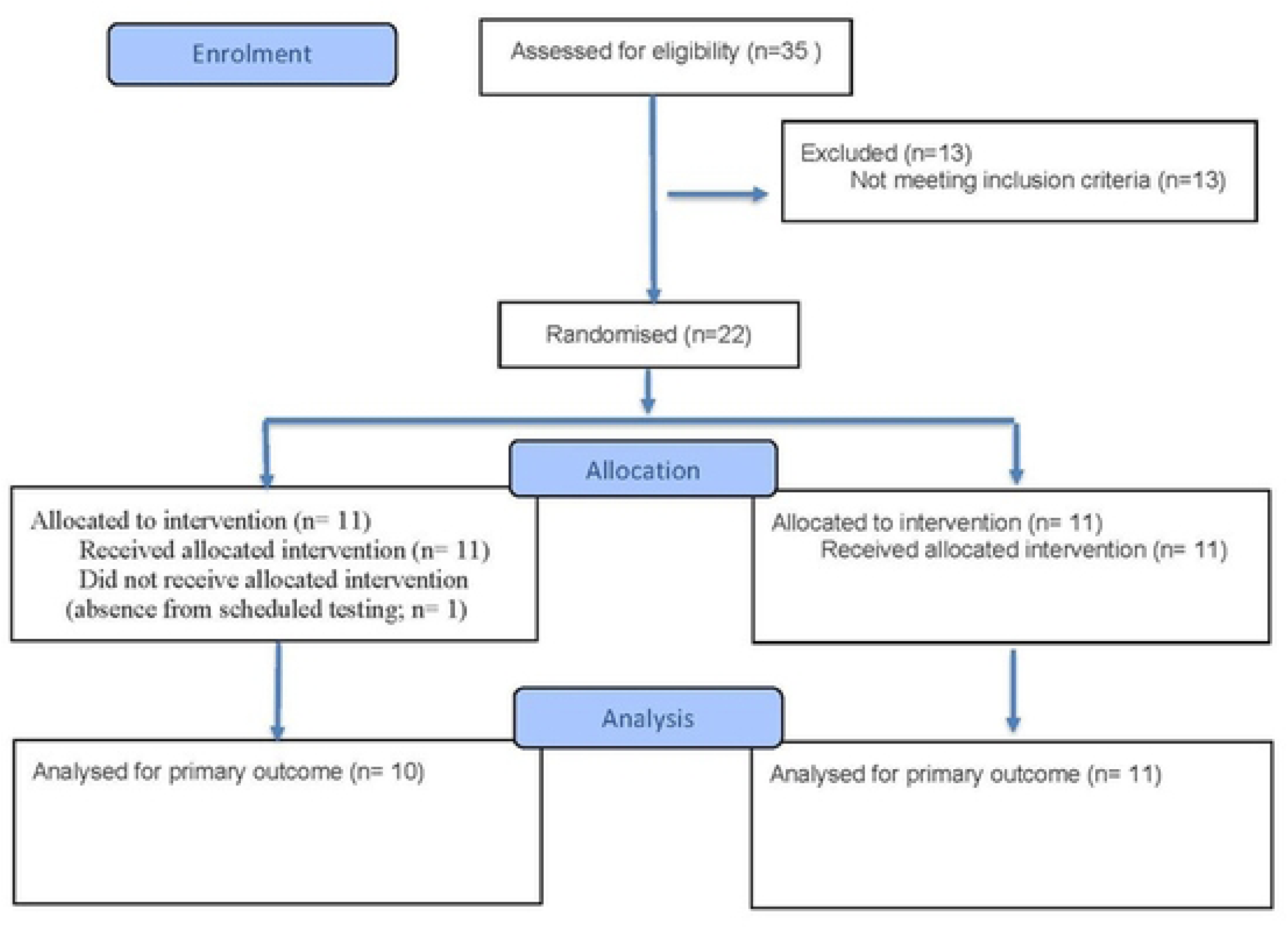
Flow Chart

This study was registered at Deutsches Register Klinischer Studien (DRKS00020280) and was approved by the local ethic committee in Bern, Switzerland (KEK- 2019-00055). This study followed the CONSORT 2010 guideline when reporting a pilot or feasibility trial (Eldridge, Chan, et al., 2016).

### Sample size

As a feasibility study, no formal sample size calculation was performed. Following CONSORT recommendations, the aim was to recruit a pragmatically defined sample sufficient to test feasibility procedures. A target range of 12 to 30 participants per group is considered appropriate for feasibility estimation, and the planned sample size aligned with this guidance.

### Recruitment period and informed consent

Participant recruitment for this prospective feasibility study took place from 01.02.2019 to 01.05.2029. All participants were healthy adults aged 20 to 35 years and were recruited voluntarily.

All participants provided written informed consent prior to inclusion. No minors were enrolled in this study, and therefore no parental or guardian consent was required.

## Randomization and Allocation

### Sequence generation

The random allocation sequence was generated using the software program “Herbers Excel/VBA Teaching and Learning Materials – The Standard Work” (Herber, 2012). A simple randomization method was applied without any restrictions such as blocking or stratification, ensuring that each participant had an equal chance of being assigned to either the intervention group or the control group.

### Allocation concealment

Group assignments were placed into sequentially numbered, opaque and sealed envelopes prepared by the statistician. Envelopes were tamper resistant and were opened only after baseline assessments were completed to ensure allocation concealment.

### Implementation

A trained research assistant enrolled participants and conducted baseline assessments without access to the randomization sequence. The study coordinator opened the next envelope in sequence to assign group allocation. The statistician who generated the sequence was not involved in enrolment, intervention delivery, or data collection, and intervention providers had no access to the allocation list.

### Blinding

Blinding was implemented wherever feasible. Outcome assessors and the statistician who performed the analyses were blinded to group allocation using anonymized datasets. Due to the nature of manual therapy, participants and therapists could not be blinded. Intervention delivery and outcome assessment were performed by separate personnel to maintain blinding integrity.

### Participants

Eligible participants were free of acute physical complaints during the four weeks preceding inclusion. Exclusion criteria included chronic pain, osteoporosis, antihypertensive medication use, acute infections, fever, fractures of the spine or pelvis within the previous 12 months, cardiac or neurological diseases, peripheral vascular disease, thrombosis, and pregnancy. Data collection took place in the research laboratory of the Bern University of Applied Sciences, School of Health Professions.

### Intervention

For the intervention group, baseline measurements were followed by six standardized manual therapy sessions over 14 days. The intervention was delivered face to face, provided individually, and conducted in a separate treatment room to ensure privacy and standardization of the setting. After assessing general well-being, participants were positioned prone on a treatment table with arms alongside the trunk and a bolster under the ankles for comfort (Fig. 2). The spinous process of C7 was identified and marked with a hypoallergenic skin marker; passive cervical extension confirmed the landmark by anterior translation of C6. The intervention was performed above T5, which was also marked. From T7, identified via the inferior scapular angles (Biel & Dorn, 1997), two spinous processes were counted cranially to locate T4 and its transverse process, approximately two fingerbreadths cranial (Reichert, 2007). Mobilization targeted the left costotransverse joint of T4 using Maitland Grade III oscillatory mobilization for 10 minutes in an anterior direction. Grade III involves large-amplitude movements at end range to increase mobility. Oscillations were performed at ∼2 Hz (≈120 movements/min) as described by Hertling and Kessler (2006). The intervention was delivered by a licensed physiotherapist with more than fifteen years of clinical experience in musculoskeletal manual therapy and formal training in Maitland mobilization techniques. The therapist had previously completed advanced postgraduate coursework in orthopaedic manual therapy and underwent a study-specific calibration session to standardize mobilization amplitude, frequency, and patient positioning. No modifications to the intervention protocol were made during the course of the study. All mobilization procedures, session frequency, duration, and positioning were delivered exactly as prespecified in the study protocol.

**Figure 2:** Mobilization techniques

### Control

The control group maintained normal daily activities and received no treatment during the 14-day period.

### Outcomes

#### Primary outcome - feasibility outcome variables

Success criteria for feasibility were defined as recruitability, adherence rate, study acceptance, and the data collection and analysis process, as reported by Rogan et al. (2016) and Tal et al. (2018). An adherence rate of more than 97% was defined a priori, as participants were young, healthy, and motivated students who provided written commitment. Likewise, the attrition rate of less than 3% was considered realistic and acceptable. In clinical practice, occasional dropouts may occur despite high patient compliance due to personal circumstances such as illness or other unforeseen events. Patient safety was assessed using provocation tests based on Carlesso et al. (2013).

**Table 1.** Outcomes of criteria of success.

|  | Assessment method | Targeted results |
| --- | --- | --- |
| Recruitability | Number of study participants (N) | N = 20 |
| Adherence | Attendance list: Measurements | Attrition rate: < 3% |
|  | T0, T1 and interventions | Adherence rate: > 97% |
|  |  | Drop-out: 0 |
| Safety | Survey using a checklist: number and severity of side effects | 0 |
| Data collection and analysis | Survey with questionnaire:<br>Number of measurement errors/inconsistent data | <5% |
|  | Duration of the investigation | 14 days |

#### Secondary outcome - physiological outcome variables

To assess the HRV, this study used the Polar Vantage V2 wristwatch, which records RR intervals in milliseconds, in line with recommendations by Sammito et al. (2014). The device has demonstrated high validity and reliability compared with ECG, with correlations of r = 0.99 (standing) and r = 0.97 (trot), and ICC values of 0.99 and 0.97, respectively (Essner et al., 2013). HRV analysis is based on RR intervals, representing the time between R-waves of the QRS complex. Measurements were performed in both time and frequency domains using spectral analysis. Key parameters included High Frequency (HF: 0.15–0.40 Hz, parasympathetic activity), Low Frequency (LF: 0.04–0.15 Hz, mixed sympathetic-parasympathetic influence), and the LF/HF ratio as an index of autonomic balance (Engel, 2010; Pagani et al., 1997).

HRV measurement: The baseline measurement was conducted at BFH in a quiet room for 10 minutes. All subsequent measurements were performed at home in a supine position: in the morning, 10 minutes after waking, and in the evening, 10 minutes before bedtime. Participants were instructed to clean the sensors before being used to minimize measurement artifacts.

### Heart rate

The Polar Vantage V2 measures not only HRV but also HR, expressed as beats per minute. This measurement was performed simultaneously with HRV recordings in the morning and evening.

Heart rate assessment: This was identical to HRV measurement.

### Data transfer and analysis

The HRV and HR data stored were transferred to the Polar Pro Trainer 5 software via infrared sensors by two blind research assistants. The data were then saved on a PC for further analysis using Kubios HRV (Standard version) developed by the Biosignal Analysis and Medical Imaging Group. Kubios HRV is widely used in HRV research and diagnostics and was employed in this study for frequency-domain analysis. Data were processed based on oscillation frequencies (ms²), and the LF/HF ratio was calculated after assigning data to respective frequency bands. Two analytical models were applied: Fast Fourier Transformation (FFT) and Autoregressive (AR) modeling. Due to reported limitations of FFT (Weippert, 2009), AR analysis was performed to validate and compare results. Final statistical analysis was conducted by an independent blinded statistician after completion of all measurements. For quality control, data entry into summary tables was performed by one researcher and verified by a second independent person.

### Statistical analyse

For this study, descriptive statistics were calculated for all measurements and presented as median, interquartile range (IQR), mean, and standard deviation (±). Within-group comparisons between baseline (T0) and post-intervention (T1) measurements were performed using the Wilcoxon signed-rank test (WSR). Subsequently, values from the intervention and control groups were compared by applying the Wilcoxon rank-sum test to assess significant differences between groups at T0 and T1. An intention-to-treat (ITT) analysis was conducted by imputing missing values with the mean of the respective parameter. Descriptive statistics and all statistical tests were performed using SPSS software (Version 28.0; SPSS Inc., Chicago, IL, USA). Statistical significance was set at p < 0.05.

## Results

A total of 22 participants were included in this study. They were randomly assigned to IG (n = 10; mean age 21.75 ± 5.36 years) or CG (n = 11; mean age 22.70 ± 2.71 years). One dropout occurred in IG due to repeated absence from scheduled testing sessions without notice. Despite follow-up attempts, the participant did not respond.

### Primary outcome - feasibility outcome variables

Two additional participants were successfully recruited; however, the predefined targets for recruitment were not achieved (Table 2). Both the attrition rate and dropout exceeded the planned thresholds, while adherence fell below the expected level. No adverse effects were observed. Data collection and analysis revealed no measurement errors. The study duration was 29 days, substantially exceeding the planned 16-day timeframe.

**Table 2.**

|  | Assessment method | Targeted results | Achieved results |
| --- | --- | --- | --- |
| Recruitability | Number of study participants (N) | N = 20 | 22 |
|  |  | (IG= 10, CG =10) | (IG= 11, CG =11) |
| Adherence | Attendance list: Measurements<br>T0, T1 and interventions | Attrition rate: < 3% | 4.5% |
|  |  | Adherence rate: > 97% | 94.5% |
|  |  | Drop-out: 0 | 1 |
| Safety | Survey using a checklist: number<br>and severity of side effects | 0 | 0 |
| Data collection and<br>analysis | Survey with questionnaire: |  |  |
|  | Number of measurement<br>errors/inconsistent data | <5% | 0% |
|  | Duration of the investigation | 16 days | 19 days |

### Secondary outcome - physiological outcome variables

Within the intervention group, large effect sizes were observed (Table 3). At the evening measurement, HF_AR showed an effect size (ES) of 0.80 with statistical significance (p = 0.008). Additionally, at the morning measurement, HF_FFT demonstrated a large ES of 0.72 (p = 0.016), HF_AR an ES of 0.78 (p = 0.010), and LF/HF_AR an ES of 0.70 (p = 0.021). No statistically significant differences were found for heart rate; however, observable trends were present (Table 4).

**Table 3.**
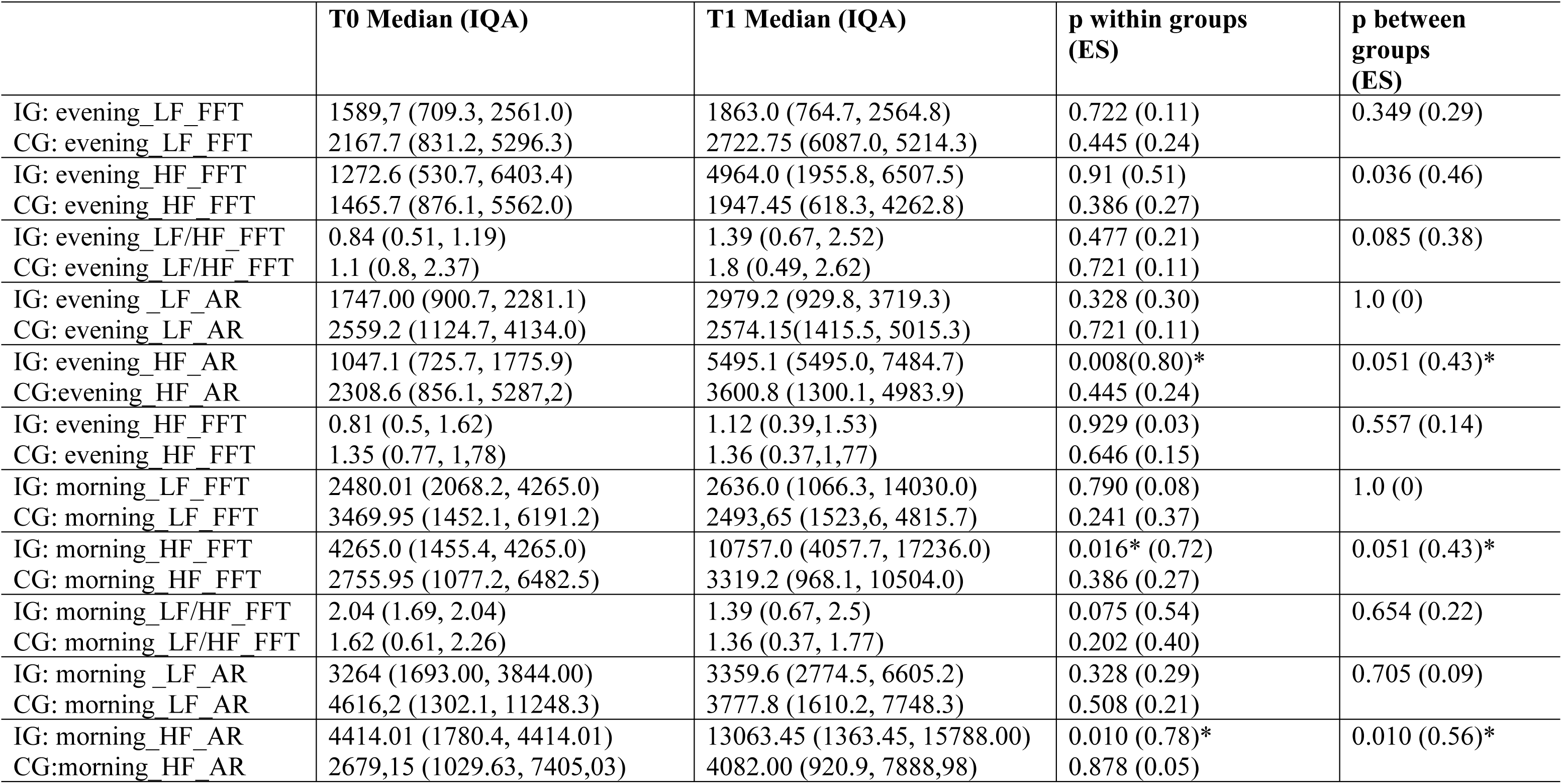
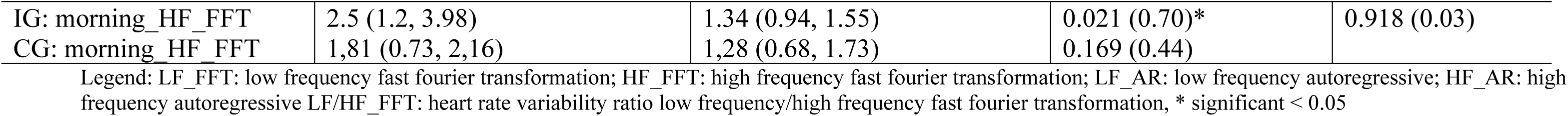
ITT results of HRV from the FFT analysis model of group 1 and group 2 in ms² (median and interquartile range IQR)

|  | **T0 Median (IQA)** | **T1 Median (IQA)** | **p within groups (ES)** | **p between groups (ES)** |
| --- | --- | --- | --- | --- |
| IG: evening_LF_FFT | 1589,7 (709.3, 2561.0) | 1863.0 (764.7, 2564.8) | 0.722 (0.11) | 0.349 (0.29) |
| CG: evening_LF_FFT | 2167.7 (831.2, 5296.3) | 2722.75 (6087.0, 5214.3) | 0.445 (0.24) |  |
| IG: evening_HF_FFT | 1272.6 (530.7, 6403.4) | 4964.0 (1955.8, 6507.5) | 0.91 (0.51) | 0.036 (0.46) |
| CG: evening_HF_FFT | 1465.7 (876.1, 5562.0) | 1947.45 (618.3, 4262.8) | 0.386 (0.27) |  |
| IG: evening_LF/HF_FFT | 0.84 (0.51, 1.19) | 1.39 (0.67, 2.52) | 0.477 (0.21) | 0.085 (0.38) |
| CG: evening_LF/HF_FFT | 1.1 (0.8, 2.37) | 1.8 (0.49, 2.62) | 0.721 (0.11) |  |
| IG: evening_LF_AR | 1747.00 (900.7, 2281.1) | 2979.2 (929.8, 3719.3) | 0.328 (0.30) | 1.0 (0) |
| CG: evening_LF_AR | 2559.2 (1124.7, 4134.0) | 2574.15(1415.5, 5015.3) | 0.721 (0.11) |  |
| IG: evening_HF_AR | 1047.1 (725.7, 1775.9) | 5495.1 (5495.0, 7484.7) | 0.008(0.80)* | 0.051 (0.43)* |
| CG:evening_HF_AR | 2308.6 (856.1, 5287,2) | 3600.8 (1300.1, 4983.9) | 0.445 (0.24) |  |
| IG: evening_HF_FFT | 0.81 (0.5, 1.62) | 1.12 (0.39,1.53) | 0.929 (0.03) | 0.557 (0.14) |
| CG: evening_HF_FFT | 1.35 (0.77, 1,78) | 1.36 (0.37,1,77) | 0.646 (0.15) |  |
| IG: morning_LF_FFT | 2480.01 (2068.2, 4265.0) | 2636.0 (1066.3, 14030.0) | 0.790 (0.08) | 1.0 (0) |
| CG: morning_LF_FFT | 3469.95 (1452.1, 6191.2) | 2493,65 (1523,6, 4815.7) | 0.241 (0.37) |  |
| IG: morning_HF_FFT | 4265.0 (1455.4, 4265.0) | 10757.0 (4057.7, 17236.0) | 0.016* (0.72) | 0.051 (0.43)* |
| CG: morning_HF_FFT | 2755.95 (1077.2, 6482.5) | 3319.2 (968.1, 10504.0) | 0.386 (0.27) |  |
| IG: morning_LF/HF_FFT | 2.04 (1.69, 2.04) | 1.39 (0.67, 2.5) | 0.075 (0.54) | 0.654 (0.22) |
| CG: morning_LF/HF_FFT | 1.62 (0.61, 2.26) | 1.36 (0.37, 1.77) | 0.202 (0.40) |  |
| IG: morning_LF_AR | 3264 (1693.00, 3844.00) | 3359.6 (2774.5, 6605.2) | 0.328 (0.29) | 0.705 (0.09) |
| CG: morning_LF_AR | 4616,2 (1302.1, 11248.3) | 3777.8 (1610.2, 7748.3) | 0.508 (0.21) |  |
| IG: morning_HF_AR | 4414.01 (1780.4, 4414.01) | 13063.45 (1363.45, 15788.00) | 0.010 (0.78)* | 0.010 (0.56)* |
| CG:morning_HF_AR | 2679,15 (1029.63, 7405,03) | 4082.00 (920.9, 7888,98) | 0.878 (0.05) |  |
| IG: morning_HF_FFT | 2.5 (1.2, 3.98) | 1.34 (0.94, 1.55) | 0.021 (0.70)* | 0.918 (0.03) |
| CG: morning_HF_FFT | 1,81 (0.73, 2,16) | 1,28 (0.68, 1.73) | 0.169 (0.44) |  |

Legend: LF\_FFT: low frequency fast fourier transformation; HF\_FFT: high frequency fast fourier transformation; LF\_AR: low frequency autoregressive; HF\_AR: high frequency autoregressive LF/HF\_FFT: heart rate variability ratio low frequency/high frequency fast fourier transformation, \* significant < 0.05

**Table 4.**
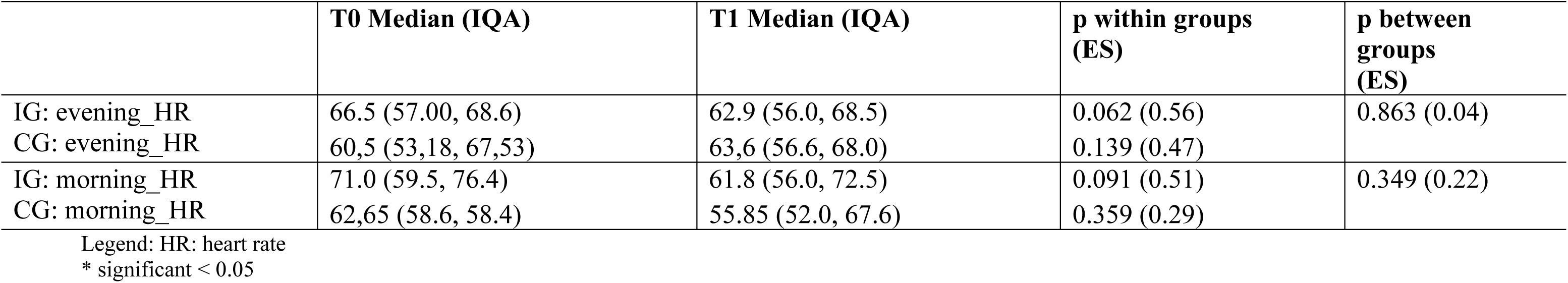
Heart rate of both groups.

|  | **T0 Median (IQA)** | **T1 Median (IQA)** | **p within groups (ES)** | **p between groups (ES)** |
| --- | --- | --- | --- | --- |
| IG: evening_HR | 66.5 (57.00, 68.6) | 62.9 (56.0, 68.5) | 0.062 (0.56) | 0.863 (0.04) |
| CG: evening_HR | 60,5 (53,18, 67,53) | 63,6 (56.6, 68.0) | 0.139 (0.47) |  |
| IG: morning_HR | 71.0 (59.5, 76.4) | 61.8 (56.0, 72.5) | 0.091 (0.51) | 0.349 (0.22) |
| CG: morning_HR | 62,65 (58.6, 58.4) | 55.85 (52.0, 67.6) | 0.359 (0.29) |  |

Legend: HR: heart rate \* significant < 0.05

## Discussion

This feasibility study primarily evaluated whether a randomized protocol for TSM can be successfully implemented in healthy participants, assessed through adherence, absence of UAE, and the practicality of acquiring and analyzing autonomic markers. The secondary aim was to provide preliminary estimates of the effects of six interventions over 14 days involving rotational mobilizations above Th5 on autonomic nervous system (ANS) outcomes, heart rate variability (HRV) and heart rate (HR) to inform the design and sample size calculation for future randomized controlled trials. Specifically, the research question was: What changes occur in HRV and related autonomic parameters following six rotational mobilizations of the thoracic spine above Th5 within 14 days?

Procedural safety and methodological integrity were achieved (no UAE; complete datasets), but feasibility was only partially met due to adherence shortfalls, higher-than-expected attrition, and device-related delays. Thus, the research question can be answered affirmatively: repeated TSM influences HRV, mainly through HF-related changes, although LF/HF interpretation remains controversial. The intervention group demonstrated large effect sizes in HRV indices: at evening assessment, HF_AR showed ES = 0.80 (p = .008); at morning, HF_FFT reached ES = 0.72 (p = .016), HF_AR ES = 0.78 (p = .010), and LF/HF_AR ES = 0.70 (p = .021). HR remained unchanged.

### Feasibility and methodological considerations

According to the decision framework proposed by Thabane et al. (2010) for interpreting feasibility study results, our predefined feasibility criteria indicated mixed performance. Safety and data completeness met the progression thresholds (“green”), whereas adherence, attrition, and recruitment fell below the predefined targets (“amber/red”). Taken together, these outcomes correspond to the category “Continue, but modify protocol, feasible with modifications.” Thus, the main trial appears feasible, provided that recruitment strategies, device logistics, and adherence enhancing measures are optimized. To enhance rigor and interpretability in future trials, improvements should include respiratory monitoring or control (e.g., paced breathing or capnography) to reduce confounding in HF and LF measures (Shaffer & Ginsberg, 2017); ECG-based HRV with rigorous artifact handling for superior timing precision (Electrophysiology, 1996), time-of-day standardization and tracking of sleep, caffeine, and activity to minimize circadian variance; multimodal autonomic readouts such as HRV, skin conductance, and blood pressure (Boucsein et al., 2012) and design rigor through preregistration, allocation concealment, and prespecified endpoints (Billman, 2013; Reyes del Paso et al., 2013). Expanding beyond healthy young adults to populations with autonomic dysregulation will improve clinical relevance (Rogan et al., 2023). In line with the CONSORT extension for pilot and feasibility trials (Eldridge, Chan, et al., 2016), we explicitly quantified our predefined progression criteria: (i) *safety* and *data completeness* met the criteria (“green”); (ii) *adherence*, *attrition*, and *recruitment within the planned timeframe* did not meet thresholds (“amber/red”). Thus, the overall judgment aligns with “continue but modify,” indicating that the protocol is feasible with operational refinements.

### Interpretation of HRV outcomes in context

In this study, the baseline HRV measurement was conducted at BFH in a quiet and standardized laboratory environment, whereas all subsequent morning and evening measurements were performed at home under less controlled conditions. This difference in measurement setting is relevant because environmental factors such as room temperature, background noise, lighting conditions, and contextual stress can influence HRV recordings. Consequently, comparisons between baseline and home-based measurements should be interpreted with caution. Environmental variability between the laboratory and the home environment may therefore have affected the autonomic outcomes, as temperature fluctuations, variable posture stability, and individual differences in the home context can modulate autonomic tone and alter HRV parameters. Although participants were instructed to standardize their measurement conditions as much as possible, residual variability cannot be completely excluded. Future studies should either use fully standardized measurement environments or incorporate environmental monitoring and documentation in order to better control for these potential confounding influences on HRV.

The HF band of short-term HRV is widely regarded as a parasympathetic (vagal) marker, provided respiration is appropriately considered (Electrophysiology, 1996; Shaffer & Ginsberg, 2017). In contrast, contemporary evidence indicates that LF, in absolute and normalized units and the LF/HF ratio do not provide a valid or specific index of cardiac sympathetic tone; LF often reflects baroreflex- and vagally mediated dynamics, and LF/HF does not quantify “sympathovagal balance” (Billman, 2013; Reyes del Paso et al., 2013). Within this framework, our HF increases are consistent with parasympathetic modulation, while the LF/HF decrease should not be overinterpreted as direct sympathetic withdrawal. The absence of significant HR changes alongside HRV modulation is plausible: HR is a coarse endpoint influenced by multiple determinants and may be less sensitive than HRV to subtle autonomic shifts in healthy, resting young adults (Electrophysiology, 1996).

Because respiration can shape HF and to a lesser extent LF, future studies should monitor or control breathing (e.g., paced breathing or capnography) to reduce confounding (Shaffer & Ginsberg, 2017). In parallel, RMSSD, a robust time-domain vagal index, should be prioritized as a primary endpoint in addition to HF(Shaffer & Ginsberg, 2017). Given current HRV standards, HF and RMSSD are the most robust short-term indices of vagal activity; future work should therefore include RMSSD as a primary time-domain endpoint alongside HF.

### Consistency and contrast with prior manual therapy research

Our findings are broadly aligned with reports of HRV modulation after manual interventions (Budgell & Polus, 2006; Jowsey & Perry, 2010), yet differ from studies showing no HRV changes (Sampath et al., 2017; Tal et al., 2018). Such discrepancies likely reflect heterogeneity in technique (mobilization vs. manipulation), body position (e.g., prone effects), dose (single vs. repeated sessions), measurement timing (morning vs. evening), and analytic methods (AR vs. FFT; respiration control). Our protocol’s repeated sessions and standardized prone positioning may have amplified mechanoreceptor input and stabilized measurement conditions, enhancing signal-to-noise relative to single-session designs.

Beyond spinal techniques, other physiotherapy modalities, e.g. massage, trigger point therapy, and self-myofascial release (foam rolling) report reductions in BP and HR and evidence of enhanced vagal activity or reduced sympathetic outflow, potentially via Ruffini-ending stimulation and nitric oxide (NO)-mediated pathways (Lastova et al., 2018; Moraska et al., 2010; Moyer et al., 2004; Schleip, 2003; Takamoto et al., 2009). In contrast, reviews focused on spinal mobilization/manipulation have observed sympathetic activation signals (e.g., changes in skin conductance/temperature) in a subset of studies (Navarro-Santana et al., 2020). A recent synthesis indicated that ANS effects occur in a majority of trials, with mixed sympathetic and parasympathetic patterns and region-independent responses (Rogan et al., 2023), consistent with a central integration perspective.

### Mechanistic plausibility: integrated autonomic patterns and vagal afferents

The present pattern fits Jänig’s view that autonomic responses arise as integrated patterns serving homeostasis, not merely reciprocal shifts (Jänig, 2022). Neuhuber (Neuhuber, 2009) emphasized the central autonomic network (CAN) and somato-autonomic reflexes, whereby thoracic mechanoreceptor input ascends through spinal and supraspinal circuits to influence brainstem and hypothalamic centers (Neuhuber, 2009). Converging neurophysiological evidence highlights midbrain structures, notably the periaqueductal gray (PAG), with monosynaptic projections to the raphe pallidus, in modulating cardiovascular and respiratory outputs (Bandler & Shipley, 1994).

Crucially, insights from Berthoud and Neuhuber (2000) on the functional and chemical anatomy of the afferent vagal system further support the plausibility that peripheral mechanosensory inputs can shape central autonomic patterning. Although the proportion of peptidergic vagal afferents is small, local effector functions via axon collateral reflexes and peptide release (e.g., CGRP, substance P) are established for dorsal root spinal afferents and suggested in select vagal contexts (Holzer, 1991, 1998; Kressel & Radespiel-Tröger, 1999). Intraganglionic laminar endings (IGLEs) exhibit complex terminal fields in the gut, including synaptic contacts with myenteric neurons, indicating close sensorimotor coupling at the enteric level (Neuhuber, 1987; Neuhuber & Clerc, 1990). Functionally, vagal-stimulation-induced gastrointestinal responses have been observed under specific pharmacological or surgical conditions (Delbro et al., 1983; Tsubomura et al., 1987), and capsaicin-sensitive afferents contribute to mucosal blood-flow regulation (Thiefin et al., 1990), although antidromic cervical vagus stimulation evokes limited c-Fos expression in myenteric neurons and fails to induce intestinal motility in some models (Neya et al., 1990; Zheng et al., 1997), underscoring context-dependent efferent/afferent interplay.

At the central level, nucleus tractus solitarii (NTS) neurons receiving primary vagal afferent input project (i) locally to dmnX, nucleus ambiguus, and RVLM, (ii) to the parabrachial/Koelliker-Fuse complex, and (iii) to forebrain sites (hypothalamic nuclei including PVN, LHA, ARC, DMH; amygdala; bed nucleus of the stria terminalis; insular cortex) (Berthoud & Neuhuber, 2000; Ter Horst et al., 1989). Given these extensive connections, vagal-afferent signals can affect organism-wide autonomic coordination, consistent with our observation of HF increases without obligatory HR changes. Historically, Lawes and colleagues proposed that vagal afferents subserve a protective role, with evolutionary roots in paraventricular pathways and defensive somatic reflexes, later elaborated into vago-vagal and vago-spinal reflexes that support homeostasis, aptly summarized as the vagus being a “great wandering protector” (Andrews & Lawes, 2020; Berthoud & Neuhuber, 2000; Leslie et al., 2020). Placed alongside PAG–raphe mechanisms and CAN organization, these vagal-afferent architectures provide a coherent explanatory frame for pattern-level modulation of HRV after repeated TSM, particularly the HF-dominant changes observed here.

### Clinical relevance and implications for future trials

Although not powered for efficacy, the large within-group effect sizes in HF (ES ≈ .72–.80) and the LF/HF change (ES ≈ .70; interpreted cautiously) provide practical inputs for sample size planning. If replicated in controlled trials with tighter physiological control and robust blinding, thoracic mobilization may be explored as an adjunct for conditions marked by autonomic imbalance, with HF/RMSSD as primary endpoints. Clinical outcomes (symptoms, function, quality of life) should accompany autonomic markers to establish translational value (Berthoud & Neuhuber, 2000; Jänig, 2022; Neuhuber, 2009).

It is also important to acknowledge that contextual factors may have contributed to the observed HRV changes. Non-specific elements such as therapist–participant interaction, touch, verbal cues, expectations, and the therapeutic setting can modulate autonomic responses independently of the specific mobilization technique. These contextual influences are increasingly recognized as active components within manual therapy mechanisms and may partially account for physiological responses observed in feasibility studies (Keter et al., 2025). In future trials, it will therefore be essential to systematically measure these contextual influences. This can be achieved by assessing treatment expectations through validated expectancy and credibility questionnaires, by quantifying the therapeutic alliance using standardized short-form instruments, and by documenting verbal and non-verbal therapist–participant communication. Additional measures such as perceived pleasantness of touch or participants’ pre-existing beliefs about manual therapy can further clarify the contribution of contextual factors. As emphasized by Keter et al. (2025), capturing these variables is crucial to disentangle specific physiological effects of mobilization from broader contextual mechanisms.

### Strengths and limitations

Strengths include a randomized protocol, safety verification, and convergent HF effects across AR and FFT at distinct time points. Limitations include restricted device availability, adherence/attrition shortfalls, lack of standardized baseline autonomic profiling, no direct respiratory control, and a healthy young adult sample that limits generalizability. Multiple HRV endpoints increase risk of Type I error in exploratory testing; accordingly, findings are hypothesis-generating (Billman, 2013; Reyes del Paso et al., 2013). Recruitment proved more challenging than anticipated, primarily because the sample consisted of young university students with fluctuating schedules, examination periods, and limited daytime availability. In addition, delays related to HRV device turnover and battery recharge cycles extended the overall study duration from the planned 16 to 29 days. These factors contributed to both reduced adherence and elevated attrition. Future trials should allocate a longer recruitment window, expand beyond a single student population, and secure redundant HRV devices to prevent bottlenecks.

## Conclusion

In line with the feasibility trial interpretation framework of Thabane et al. (2010), feasibility was partially achieved, corresponding to “continue but modify protocol.” Safety and data completeness criteria were fully met, whereas predefined thresholds for adherence (>97%), attrition (<3%), and recruitment (N=20 within 16 days) were not achieved. According to the CONSORT progression criteria for pilot/feasibility studies (Eldridge, Chan, et al., 2016), safety and data completeness were met, while adherence and timely recruitment were unmet. Physiologically, repeated TSM produced consistent increases in HF-HRV, suggesting vagal modulation, whereas LF/HF should be interpreted cautiously. Future confirmatory trials should incorporate respiratory control, prioritize HF and RMSSD as primary vagal indices, use ECG-based HRV, and address logistical barriers (e.g., device availability, delayed scheduling). The observed effect sizes (ES ≈ .70–.80) may support sample size calculation for future fully powered trials.

## Data Availability

Data are available from correspondending author

